# Single-exon deletions of *ZNRF3* exon 2 cause congenital adrenal hypoplasia

**DOI:** 10.1101/2023.06.14.23290643

**Authors:** Naoko Amano, Satoshi Narumi, Katsuya Aizu, Mari Miyazawa, Kohji Okamura, Hirofumi Ohashi, Noriyuki Katsumata, Tomohiro Ishii, Tomonobu Hasegawa

**Author notes:** **Correspondence:** Tomonobu Hasegawa, M.D., Ph.D.

## Abstract

Primary adrenal insufficiency (PAI) is a life-threatening condition characterized by the inability of the adrenal cortex to produce sufficient steroid hormones. E3 ubiquitin protein ligase zinc and ring finger 3 (ZNRF3) is a negative regulator of Wnt/β-catenin signaling. R-spondin 1 (RSPO1) enhances Wnt/β-catenin signaling via binding and removal of ZNRF3 from the cell surface. To explore a novel genetic form of PAI, we analyzed nine childhood-onset patients with PAI of biochemically and genetically unknown etiology using array comparative genomic hybridization. We identified various-sized single-exon deletions encompassing of *ZNRF3* exon 2 in three patients, who showed neonatal-onset adrenal hypoplasia with glucocorticoid and mineralocorticoid deficiencies. RT-PCR analysis showed that the three distinct single exon deletions were commonly transcribed into a 126-nucleotide deleted mRNA and translated into 42-amino acid deleted protein (ΔEx2-ZNRF3). Three-dimensional structure modeling predicted that interaction between ZNRF3 and RSPO1 was disturbed in ΔEx2-ZNRF3, suggesting loss of RSPO1-dependent activation of Wnt/β-catenin signaling. Cell-based functional assays with the TCF-LEF reporter showed that RSPO1-dependent activation of Wnt/β-catenin signaling was attenuated in cells expressing ΔEx2-ZNRF3 as compared with those expressing wild-type ZNRF3. In summary, we provided genetic evidence linking deletions encompassing ZNRF3 exon 2 and congenital adrenal hypoplasia, which might be related to constitutive inactivation of Wnt/β-catenin signaling by ΔEx2-ZNRF3.

## Introduction

Primary adrenal insufficiency (PAI) is a life-threatening condition characterized by the inability of the adrenal cortex to produce sufficient glucocorticoids and/or mineralocorticoids. The major etiology of childhood-onset PAI is congenital adrenal hyperplasia such as 21-hydroxylase deficiency^1^, which shows specific biochemical features including a high serum level of 17α-hydroxyprogesterone. A subset of patients with childhood-onset PAI lack these biochemical features and need genetic testing for the etiological diagnosis. Historically, eight causative genes for biochemically uncharacterized PAI (*AAAS, CYP11A1, MC2R, MRAP, NR0B1, NR5A1* and *STAR*) have been identified by 2010^2–8^. In the 2010s, the introduction of whole exome sequencing (WES) into genetic analysis for PAI led the identification of additional causative genes, including *NNT, CDKN1C, MCM4, SAMD9*, *SGPL1* and *TXNRD2*^9–13^. Now, the etiologies of the majority patients with biochemically uncharacterized PAI can be elucidated^14–17^, however those of the rest remains unknown.

In the Wnt/β-catenin signaling, binding of Wnt ligands to Frizzled receptors on the cell membrane results in translocation of β-catenin into the nucleus and expression of specific target genes. E3 ubiquitin protein ligase zinc and ring finger 3 (ZNRF3) acts as a negative regulator of Wnt/β-catenin signaling by targeting Frizzled receptors for proteasomal degradation^18^. R-spondin 1 (RSPO1) binds the extracellular domain of ZNRF3 and promotes its removal from the cell surface, which ultimately enhances Wnt/β-catenin signaling (Figure S1A,B). In the process of zonation of the adrenal cortex in mice, the expression level of β-catenin is high in progenitor cells localized on a subcapsular region and gradually decreases from subcapsular zone to zona fasciculata^19^. Adrenocortical-specific β-catenin knockout mice develop adrenal aplasia, indicating the essential role of Wnt/β-catenin signaling in adrenal organogenesis^19^. Although the subcellular localization of β-catenin in human fetal and postnatal adrenal cortex was demonstrated^20^, the role of Wnt/β-catenin signaling in adrenocortical development remains unclear. As the relation of Wnt/β-catenin signaling to human adrenal cortex, it is known that constitutive activation of Wnt/β-catenin pathway is a major feature of adrenocortical carcinoma. Loss-of-function mutations of *ZNRF3* are one of the frequent somatic molecular alterations of adrenocortical carcinoma, found in about 20% of the specimens^21, 22^. These lines of evidence suggest the role of the Wnt/β-catenin pathway in regulation of growth of human adrenocortical cells.

Here, we report the identification and functional characterization of genomic deletions encompassing exon 2 of *ZNRF3*, which were associated with congenital adrenal hypoplasia.

## Methods

### Patients

We collected genomic DNA samples from 63 childhood-onset patients with biochemically uncharacterized PAI at Department of Pediatrics, Keio University School of Medicine. The patients were diagnosed as having PAI based on symptoms and endocrinological data (low or normal serum cortisol levels with high plasma adrenocorticotropic hormone levels) at age less than 15 years. We evaluated all the patients with urine or serum steroid profiling and confirmed that none had findings compatible with known enzymatic defects including 21-hydroxylase deficiency, P450 oxidoreductase deficiency^23, 24^. For the 63 patients, 12 known PAI-related genes (*AAAS, CDKN1C, CYP11A1, MCM4, MC2R, MRAP, NNT, NR0B1, NR5A1, SAMD9, STAR* and *TXNRD2*) were sequenced as previously described, and genetic diagnosis was determined in 54 samples^16^. Out of the remaining nine patients, eight patients had sufficient amounts of the samples, and were enrolled to the present study. We also obtained another DNA sample derived from PAI of biochemically and genetically unknown etiology that was collected at National Research Institute for Child Health and Development.

### CNV analysis

Screening for copy number variations (CNVs) was performed with oligonucleotide array comparative genomic hybridization (aCGH): Catalog arrays (SurePrint G3 Human 1M or 180K; Agilent Technologies, Santa Clara, CA) or a custom-made 60K array that covered 158-kb region corresponding to *ZNRF3* introns 1 to 2 (hg38 Chr22: 28,884,067-29,042,494) with 2,146 probes (Agilent Technologies) were used. For confirmation of the genomic deletion, fluorescent in situ hybridization (FISH) analyses were performed on cultured lymphoblasts established from P1 and P2 as well as their parents. The 9-kb FISH probe was created by long PCR of the bacterial artificial chromosome probe RP11-480L23 (Advanced Geno Techs Co., Ibaraki, Japan). The primer sequences were summarized in Table S1.

For P1 and P3, deletion breakpoints were determined by PCR and sequencing using breakpoint-specific primer pairs (Table S1). For P2, the deletion breakpoint was analyzed with whole genome sequencing using TruSeq DNA PCR-Free Library Prep Kit (Illumina, San Diego, CA) and a HiSeq 2000 sequencer (Illumina). The obtained reads were mapped to the human reference sequence GRCh37 with decoy sequences (hs37d5) using the MEM algorithm of BWA^25^. The contig sequences covering the putative breakpoint were constructed using GrepWalk as previously described^26^. The sequence around the breakpoints was also confirmed by PCR-based sequencing with a breakpoint-specific primers (Table S1). Allele frequencies of the deletions encompassing exon 2 of *ZNRF3* as identified in this study were calculated using the two databases of gnomAD SVs v2.1^27^ and jMorp^28^.

### RT-PCR

For P1 and P2, we established Epstein-Barr virus-transformed lymphoblastoid cell lines. Total RNA was isolated from the cell lines using NucleoSpin RNA (Takara Bio, Shiga, Japan). The RNA was reverse transcribed into cDNA using PrimeScript II High Fidelity RT-PCR Kit (Takara) and oligo-dT, which was followed by PCR amplification, subcloning and sequencing with primers spanning *ZNRF3* exons 1 to 5 (Figure 1A; Table S1). For P3, we assessed the effect of the deletion on mRNA with *in vitro* experiments, because living cells were unavailable. We generated HEK293 cells with 9.5-kb deletion (hg38 Chr22: 2 8,985,945-28,995,398), which included the 8.1-kb deleted region (hg38 Chr22: 28,986,799-28,994,878) identified in P3, using CRISPR/Cas9-based genome editing. Two guide RNAs (gRNAs) sequences that flank *ZNRF3* exon 2 (Figure S2A) were cloned into pX260-U6-DR-BB-DR-Cbh-NLS-hSpCas9-NLS-H1-shorttracr-PGK-puro (Addgene #42229). The sequences of gRNA1 and 2 were described in Table S1. HEK293 cells cultured in a 12-well plate were transfected with 0.5 μg of each CRISPR/Cas9 construct (gRNA1 and gRNA2) using Lipofectamine 2000 (Thermo Fisher Scientific, Waltham, MA). Forty-eight hours after transfection, puromycin (5 μg/mL) was added to the culture media to enrich the cells expressing the plasmid vectors. The selected cells were collected at 96 hours after transfection, and genomic DNA was extracted and sequenced to confirm the genome editing. RT-PCR sequencing of the *ZNRF3* mRNA was performed as described above.

**Figure 1.**
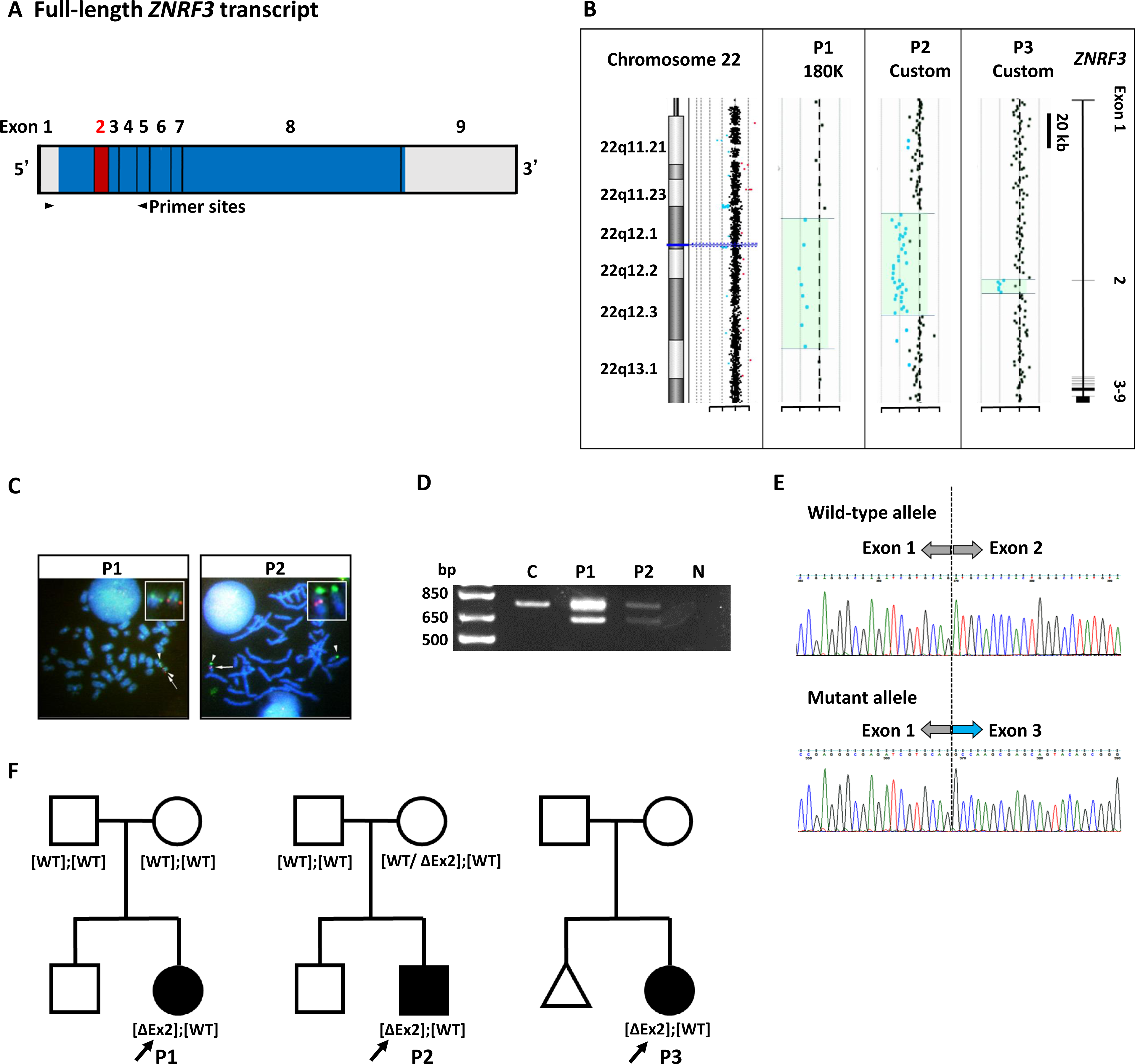
Identification of exon 2 deletions in *ZNRF3* in three PAI patients using CNV analyses and RT-PCR. **A.** Schematic diagram of full-length *ZNRF3* transcript. Blue and red areas represent translated region, gray untranslated region, and red exon 2. The facing arrowheads indicate primer sites for RT-PCR. **B.** Array CGH plots for P1-3, who have various-sized deletions encompassing exon 2 in *ZNRF3*. On the left panel, blue line indicates the region of interest (*ZNRF3*) in an idiogram of 22q11.2-22q13.1. Green-shaded region decreased in copy number, and unshaded region had normal copy number. Right panel shows the gene structure of *ZNRF3*. **C.** FISH study in P1 and P2, who have exon 2 deletions in *ZNRF3*. Both patients have one red signal (arrow) on the region of *ZNRF3* exon 2 on Chr22, and two green control signals (arrow heads) on a pair of Chr22. **D.** Agarose-gel electrophoresis of RT-PCR amplicons using the primers spanning *ZNRF3* exons 1 to 5 (Figure 1A). P1 and P2 have two fragments of 759- and 633-bp, and control (C) has the only 759-nt fragment. E. Sequence chromatograms of the obtained RT-PCR amplicons from P1 and P2. P1 and P2 have both a wild-type allele and a mutant allele skipping exon 2 sequence. F. Pedigrees of three families with exon 2 deletions in *ZNRF3*. Solid symbols denote affected individuals with neonatal-onset PAI, open symbols unaffected ones. WT and ΔEx2 denote wild-type and exon 2 deletion, respectively. Maternal mosaicism of ΔEx2 was identified in P2 family.

### Three-dimensional structures

The three-dimensional complex structure of mouse Znrf3 and human RSPO1 was visualized using PDB atomic coordinate files (4CDK) and PyMOL version 0.99.

### Functional characterization of the mutant ZNRF3

A vector encoding the partial ZNRF3 cDNA sequence (GenBank accession# NM_001206998) corresponding to exons 2 to 9 (pF1KSDA1133) was purchased from Kazusa DNA Research Institute (Chiba, Japan). The cDNA sequence corresponding to exon 1 was PCR amplified from a genomic DNA sample. Using restriction endonucleases, the two pieces of cDNA sequence were cloned into pEGFP-N1 (Takara Bio) with deleting the EGFP cDNA sequence. An expression vector for the exon 2 deleted (ΔEx2)-ZNRF3 was created by replacing the WT sequence around exon 2 with chemically-synthesized DNA (Eurofin Genomics, Tokyo, Japan) lacking the exon 2 sequence. We also created 3xFLAG-tagged ZNRF3 expression vectors by inserting chemically synthesized tag sequence between Ala55 and Lys56, where the sequence for signal peptide ends. A previously described constitutive active artificial variant p.His102Ala;Pro103Ala (H102A/P103A) was generated with standard site-directed mutagenesis technique (QuikChange II Site-Directed Mutagenesis Kit, Agilent Technologies) ^29^. These *ZNRF3* cDNA (WT, ΔEx2, and H102A/P103A mutant) were cloned into pB513B-1 (System Biosciences, Palo Alto, CA). The sequences of all constructs were verified by Sanger sequencing.

For immunodetection studies, HEK293 cells were transiently transfected with each 3xFLAG-ZNRF3 vector using Lipofectamine 2000, and the expressed proteins were detected with anti-FLAG M2 monoclonal antibody (Sigma-Aldrich, St. Louis, MO) used at 500:1 dilution. Western blotting was performed with whole cell lysates prepared from the transfected cells. Immunofluorescence analysis was performed by fixing transfected cells with 4% paraformaldehyde in phosphate-buffered saline (PBS), blocking with 1% bovine serum albumin in PBS, and incubation with the primary antibody and a secondary antibody (Alexa Fluor 555-conjugated goat anti-mouse IgG antibody, Thermo Fisher Scientific). Cells were observed under a TCS-SP5 confocal microscope (Leica Microsystems, Tokyo, Japan).

For luciferase reporter assays, stable HEK293 cell lines expressing each ZNRF3 protein (WT, ΔEx2, or H102A/P103A) were established with the piggyBac vectors according to the manufacturer’s protocol. The stable cells grown in a 96-well plate were transiently transfected with a reporter vector (luc2P/TCF-LEF-RE; pGL4.49, Promega, Madison, WI), using Lipofectamine 2000. Forty-eight hours after transfection, the cells were incubated with 0.6 mg/L of human Wnt3a (R&D Systems, Minneapolis, MN) and/or 10 mg/L of human RSPO1 (R&D Systems), in DMEM supplemented with 10% fetal bovine serum for 24 hours at 37°C. Luciferase assays were performed using ONE-Glo Luciferase Assay System (Promega) according to the manufacturer’s instructions.

### In situ hybridization of Znrf3, Shh and Nr5a1 in mouse adrenals

A 652-bp DNA fragment corresponding to the cDNA nucleotide position 5,125-5,776 of mouse Znrf3 (GenBank accession# NM_001080924.2) was subcloned into pGEMT-Easy vector (Promega). Sense and anti-sense RNA probes were generated using the vector. In the same way, sense and anti-sense RNA probes of mouse Shh (365 nt, nucleotide position 1796-2160, accession# NM_009170) and mouse Nr5a1 (586 nt, nucleotide position 1,625-2,210, accession# NM_139051.3) were generated. The sections at 6-µm thickness of paraffin-embedded mouse adrenal (C57BL/6 mouse, male, E16.5, Sankyo Labo service, Tokyo, Japan) were hybridized with digoxigenin labeled RNA probes at 60℃ for 16 hr. The bound label was detected using NBT-BCIP solution (Sigma-Aldrich), an alkaline phosphate color substrate. The sections were counterstained with Kernechtrot (Muto Pure Chemicals Co. Ltd., Tokyo, Japan)

### Study approval

We obtained written informed consent for study participation from parents. The Ethics Committee of Keio University School of Medicine approved the study (#20170130).

## Results

### Patient characteristics

For this study, we enrolled nine Japanese patients with childhood-onset PAI of biochemically and genetically unknown etiology. Seven patients were diagnosed as PAI during the neonatal period, one during infancy, and one during childhood. Seven patients were males, and the other two female. Six of the nine patients received mineralocorticoid replacement therapy for salt-wasting symptoms or persistently high plasma renin activity. All the patients did not have a family history of PAI or unexplained death in childhood. Three patients had extra-adrenal symptoms, including two with ventricular septal defect (VSD) and one with intrahepatic bile duct hypoplasia.

### Identification of ZNRF3 exon 2 deletions

For the three patients with extra-adrenal symptoms, we considered contiguous gene deletion syndrome as a possible mechanism. To screen such abnormalities, we performed aCGH in the three patients (P1-3). Although we did not observe any CNVs involving two or more genes, we unexpectedly found that two of them (P1 and P2) had heterozygous single-exon deletions encompassing exon 2 of *ZNRF3* (Figure 1B). The presence of the deletions was confirmed with FISH analysis (Figure 1C). The deletion in P1 occurred *de novo*, while the deletion in P2 was transmitted from unaffected mother with somatic mosaicism: FISH analysis of the unaffected mother showed 5 out of 25 cells had the deletion (data not shown). We then screened *ZNRF3* microdeletion in the remaining 7 patients with custom-made tiling aCGH (average probe spacing of 74 bp) and found another patient (P3) with an 8-kb deletion involving *ZNRF3* exon 2 (Figure 1B). Breakpoints of the three deletions were determined with deletion-specific PCR or whole genome sequencing (Figure S1). The sizes of deletion in P1, P2 and P3 were 73.5 kb, 59.1 kb and 8.1 kb, respectively. Deletions encompassing *ZNRF3* exon 2 were not observed in 21,694 alleles analyzed in the gnomAD SVs v2.1 and in 16,600 alleles analyzed in the 8.3KJPN-SV of jMorp. The deletion breakpoints involved *Alu* sequence in P1 and P2, but not in P3 (Figure S1). For P2, additional sequence of poly (A) and (hg38 Chr10: 99,837,467-99,840,412 or Chr19: 29,898,334-29,901,988) was inserted between the breakpoints.

The effect of exon 2 deletions on the ZNRF3 mRNA sequence was tested by RT-PCR using patient-derived lymphoblastic cell lines in P1 and 2, showing a skip of exon 2 with 126-nt length (Figure 1D, 1E). For P3, whose living cell materials were unavailable, we recapitulated the deletion in HEK293 cells with CRISPR/Cas9 technology and confirmed the presence of the identical 126-nt deleted mRNA in the cells (Figure S2B).

### Clinical findings of patients with ZNRF3 exon 2 deletion

All three patients (two males and one female) had neonatal-onset PAI without any family history of PAI or unexplained death (Table 1; Figure 1F). They had high plasma renin activity, indicating mineralocorticoid deficiency. Adrenal imaging (computed tomography, ultrasonography, or magnetic resonance imaging) showed adrenal hypoplasia (data not shown). P1 and P2 had VSD. Surgical repair of VSD was performed in P1, while spontaneous closure of VSD was seen in P2. P1 and P2 showed normal growth and pubertal development. Clinical information of P3 was unavailable except for the neonatal period.

**Table 1.** Clinical phenotype of patients with exon 2 deletion in *ZNRF3*.

| Patient | 1 | 2 | 3 |
| --- | --- | --- | --- |
| Sex | Female | Male | Male |
| Gestational age (week) | 39 | 41 | 40 |
| *ZNRF2* exon 2 deletion | Chr22:<br>28,952,131-29,025,672 | Chr22:<br>28,948,178-29,007,259 | Chr22:<br>28,986,799-28,994,878 |
| Birth weight (SDS) | -2.0 | 1.4 | -1.8 |
| Age at diagnosis of PAI | Neonatal period | Neonatal period | Neonatal period |
| Presenting symptom | Hyperpigmentation,<br>convulsion | Hyperpigmentation,<br>vomiting | Hyperpigmentation,<br>failure to thrive |
| ACTH (pg/mL)<br>(RR 25-100) | 3,840 | 2,193 | 1,240 |
| Cortisol (μg/dL)<br>(RR 5-23) | <1.0 | 2.5 | 11.1 |
| PRA (ng/mL/hr)<br>(RR 0.47-3.13) | 361.8 | 52.5 | 11.9 |
| Aldosterone (pg/mL)<br>(RR 50-900) | 51 | 3 | 24.1 |
| Adrenal imaging | Hypoplasia by CT | Hypoplasia by US and<br>MRI | Not swelling by MRI |
| Age at last visit (yr) | 10-15 | 10-15 | <1 |
| Height/Weight (SDS) | -2.0/-1.4 | -0.6/-0.3 | -2.8/-1.7 |
| Pubertal development | Breast development | Testicular volume 8mL | NA |
| LH/FSH (mIU/mL) | 0.34/3.30 | 1.03/3.48 | NE |
| Sex hormone level | E<sub>2</sub> 29.6 pg/mL | T 379 ng/dL | NE |
| Additional features | VSD (closure surgery<br>less than age of 1yr),<br>epidural hematoma | VSD (spontaneous<br>closure) | None |

Abbreviations: ACTH, adrenocorticotrophic hormone; CT, computed tomography; E2, estradiol; FSH, follicular stimulating hormone; LH, luteinizing hormone; MRI, magnetic resonance imaging; NA, not applicable; NE not examined; PAI, primary adrenal insufficiency; PRA, plasma renin activity; RR, reference range; SDS, standard deviation score; T, testosterone; US, ultrasonography; VSD, ventricular septal defect.

### Three-dimensional modeling of Znrf3-RSPO1

ZNRF3 is a single-transmembrane E3 ubiquitin ligase, which contains N-terminal signal sequence, a single pass transmembrane domain, and an intracellular C-terminal RING domain. Since the sequences of the extracellular region of human ZNRF3 and murine Znrf3 are identical except for 2 residues (Tyr^77^ and Thr^91^ in human), we used co-crystal structure data of murine Znrf3-human RSPO1 to gain structural insights into the effect of the exon 2 deletion (Figure 2A). The extracellular region of Znrf3/ZNRF3 consists of 166 amino acid (aa) residues (54-219 aa) and corresponds to exons 1 to 4. Binding to RSPO1 occurs at six residues, Gln^100^, Met^101^, His^102^, Tyr^116^, Lys^125^ and Glu^127^, of which five residues except Gln^100^ are encoded by exon 2^31^. Therefore, we predicted that the deletion of the residues encoded by exon 2 causes loss of binding to RSPO1.

**Figure 2.**
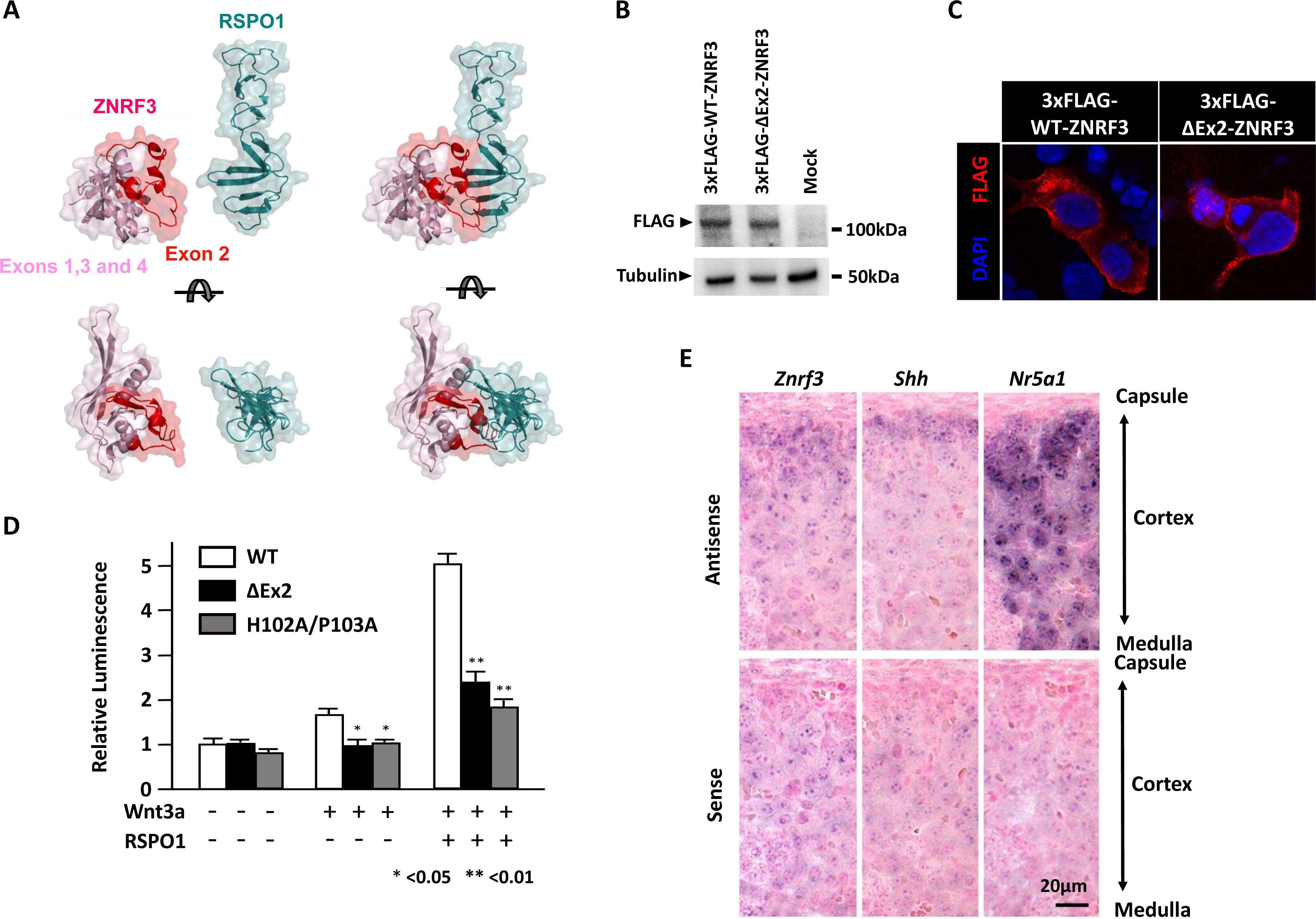
Functional characterization of exon 2 deletion in *ZNRF3*. **A.** 3D structure of Znrf3-RSPO1 complex. Transparent surface representation of the extracellular region of Znrf3 (pink; Exon 1,3 and 4, red; Exon 2) and RSPO1 (green). Based on the two views, exon 2 of Znrf3 makes a contact to RSPO1. **B.** ZNRF3 protein expression analyzed by means of Western blotting. WT and ΔEx2 protein expression in transfected HEK293 cells are comparable. **C.** Intracellular localization by immunofluorescence staining of transfected HEK293 cells with WT and ΔEx2 constructs. WT and ΔEx2 FLAG-tagged ZNRF3 (red) are localized to the plasma membrane. DAPI (blue) shows cell nuclei. **D.** The relative effect of each ZNRF3 protein (WT, ΔEx2, or H102A/P103A) on Wnt/dependent β-catenin signaling, as assessed by means of a luc2P/TCF-LEF-reporter assay. H102A/P103A denotes the previously described constitutive active artificial variant of p.His102Ala;Pro103Ala (34). The luciferase activity under RSPO1 and Wnt3a treatment was lower in cells expressing ΔEx2, as well as of H102A/P103A mutant, than those expressing WT. Data are representative of three independent experiments. E. Expression of *Znrf3, Shh and Nr5a1* in adrenal cortex of fetal mice (E16.5) by means of in situ hybridization (ISH). Upper panels show ISH with antisense probes, and lower panels ISH with sense probes. *Znrf3*, as well as *Shh*, mainly expressed in subcapsular cells of the adrenal cortex, while *Nr5a1* was expressed throughout the adrenal cortex.

### Functional characterization of the mutant ZNRF3

To examine the functionality of the exon 2 deletion (ΔEx2)-ZNRF3 protein, we performed *in vitro* functional studies with HEK293 cells. Western blotting showed a comparable protein expression level of ΔEx2-ZNRF3 with wild-type (WT)-ZNRF3 (Figure 2B, Figure S4). Immunofluorescence studies under nonpermeabilized condition showed that both ΔEx2-ZNRF3 and WT-ZNRF3 expressed on the plasma membrane (Figure 2C). The ability of each ZNRF3 protein (WT, ΔEx2, or H102A/P103A) to attenuate Wnt/dependent β-catenin signaling was tested with luc2P/TCF-LEF reporter. The luciferase activity under RSPO1 and Wnt3a treatment was significantly lower in cells expressing ΔEx2-ZNRF3, as well as of previously reported RSPO1-binding defective H102A/P103A-ZNRF3, than those expressing WT-ZNRF3 (Figure 2D).

### Expression of Znrf3 in mouse adrenals

For localizing Znrf3 expression in mouse adrenal glands at E16.5, we performed *in situ* hybridization using RNA probes against mRNAs of *Znrf3*, *Shh*, and *Nr5a1* (Figure 2E). *Znrf3* mainly expressed in subcapsular cells of the adrenal cortex as well as *Shh*, while *Nr5a1* was expressed throughout the adrenal cortex.

## Discussion

In this study, we identified various-sized heterozygous deletions (73.5, 59.1 and 8.1 kb) commonly encompassing *ZNRF3* exon 2 in three patients, who were diagnosed as having neonatal-onset adrenal hypoplasia with glucocorticoid and mineralocorticoid deficiencies. The two teenaged patients (P1 and 2) seemed to have normal gonadal function. The deletion in P1 occurred *de novo*, while it was transmitted to P2 from the mother, who had the deletion in a somatic mosaic state. No deletions involving *ZNRF3* exon 2 are registered in the gnomAD SV v2.1 and jMorp (8.3KJPN-SV) that analyzed more than 19,000 subjects. These lines of genetic evidence indicate that heterozygous deletions of *ZNRF3* exon 2 were causally related to a novel genetic form of congenital adrenal hypoplasia.

We presume that deletion of the exon 2-derived 42 amino acid residues would impair the binding to RSPO1 based on the three-dimensional modeling of the RSPO1-Znrf3 complex. This impairment would allow ZNRF3 to escape RSPO1-dependent clearance, resulting in constitutional inactivation of the Wnt-β-catenin signaling pathway (Figure S1C). The results of our *in vitro* functional studies were consistent with this presumption. This dominant-negative nature would explain the reason for causative relation of heterozygous deletions of *ZNRF3* exon 2 to congenital adrenal hypoplasia.

Adrenocortical-specific *Znrf3* knockout mice show adrenocortical hyperplasia ^32^. In these mice, the gradient of β-catenin expression levels according to the adrenocortical zones is disrupted, and β-catenin level in zona fasciculata is abnormally high as well as in subcapsular zone and zona glomerulosa. In our patients with *ZNRF3* exon 2 deletion, contrastingly, Wnt/β-catenin signaling is presumably suppressed in progenitor cells of subcapsular zone, and this would result in disturbance in the process of adrenocortical zonation. It is also worth to note that experimentally verified loss-of-function *ZNRF3* variants (p.Ser554Asn and p.Arg768Gly) have been found in patients with 46,XY disorders of sex development ^33^. These clinical observations do not contradict our model that the loss of interaction of ZNRF3 with RSPO1 maintains its inhibitory activity against Wnt/β-catenin signaling and ultimately leads to congenital adrenal hypoplasia.

Exon 2 deletions of *ZNRF3* have not been reported as a causative genetic abnormality for congenital adrenal hypoplasia even after the introduction of WES. This may be because it is generally difficult to identify single-exon deletions by WES, as well as aCGH. Several algorithms have been developed to identify CNVs from exome data, but they have not achieved sufficient accuracy for CNVs of less than three consecutive exons ^34^. Our experience suggests that the introduction of sensitive methods for detecting single-exon CNVs, such as whole genome sequencing and optical genome mapping ^35^, may identify new mechanisms involved in congenital diseases, including adrenal hypoplasia.

In our study, two patients with relatively large deletions (P1 and P2; 73.5 and 59.1 kb) had VSD, while P3 carrying a small deletion (8.1 kb) did not. VSD is thought to be a multifactorial disorder, and its prevalence (1.76-4.48 in 1,000) ^36^ is much higher than that of PAI ^37^. At this time, it is difficult to conclude whether *ZNRF3* exon 2 deletion is responsible for a novel autosomal dominant form of VSD. Genome-wide investigations of CNVs by aCGH showed that the rare CNVs identified in 12% of non-syndromic CHD patients were enriched for genes associated with the Wnt/β-catenin signaling pathway, implicating that the alteration of this signaling could increase the susceptibility to the development of non-syndromic CHD ^38^. Further accumulation of cases would be necessary to elucidate the association between *ZNRF3* exon 2 deletions and VSD.

In conclusion, we have screened CNVs in patients with childhood-onset PAI of unknown etiology and identified three patients with heterozygous deletions encompassing *ZNRF3* exon 2. Those patients showed neonatal-onset adrenal hypoplasia with glucocorticoid and mineralocorticoid deficiencies. We performed expression experiments and showed the constitutional inactivation of Wnt/β-catenin signaling pathway by the ΔEx2-ZNRF3 protein *in vitro*. These observations indicate that single exon deletions encompassing *ZNRF3* exon 2 cause congenital adrenal hypoplasia. Our investigations provide novel biological evidence that Wnt/β-catenin signaling plays a critical role for the development of human adrenal cortex.

## Data Availability

All data produced in the present work are contained in the manuscript

## Acknowledgement

We thank the physicians who referred patients to us and the patients and families for their involvement. This work was funded in part by the grant from the Ministry of Health, Labor and Welfare of Japan (20FC1020 to T.I., T.H., and N.A. for research on intractable diseases). N.A. was supported by the grants from the Japan Society for the Promotion of Science (26893259, 16K09998, and 19K08260). T.I. was supported by the grants from the Japan Society for the Promotion of Science (22K08680).

## Author contributions

N.A., S.N., T.I., and T.H. planned the study. K.A., M.M., and N.K. collected the patients’ samples and clinical information. N.A. and S.N. performed molecular analyses and N.A. performed the other experiments and analyses. K.O. performed the bioinformatic analysis for identification of the deletion breakpoint using whole genome sequencing, and H.O. established the lymphoblasts. N.A. and S.N. wrote the manuscript in consultation with T.I. and T.H.

## Web resources

gnomAD SVs v2.1, https://gnomad.broadinstitute.org/

jMorp, https://jmorp.megabank.tohoku.ac.jp/

PDB, https://www.rcsb.org/

PyMOL version 0.99, http://www.pymol.org/

