## Supplemental file for "Single-exon deletions of *ZNRF3* exon 2 cause congenital adrenal hypoplasia"

**Table S1. The list of sequences used in this study**

| Name | Sequence |
|---|---|
| Primer for FISH probe-F | 5’-GAG AAG GAC CCA TAG AAT CTG CTG GAA C-3’ |
| Primer for FISH probe-R | 5’-AAC ATC TGG CCC AAG GGT GTA AGA TAA A-3’ |
| Primer for the deletion breakpoints of P1-F | 5’-TAT CTG GCT TTC TTT TCT CCG AGT G-3’ |
| Primer for the deletion breakpoints of P1-R | 5’-GAA GGA GAA AAG TGG CAG AGT CAT C-3’ |
| Primer for the deletion breakpoints of P3-F | 5’-TGA AGT CTA GAA GCC AAA CAA TCT TGG A-3’ |
| Primer for the deletion breakpoints of P3-R | 5’-AGT AAG TAG AAA ATT TCC CCA GGG CAT C-3’ |
| Primer for the deletion breakpoints of P2-F | 5’-ATC CGC CCA CCT TGG CCT CCC AAA GTG-3’ |
| Primer for the deletion breakpoints of P2-R | 5’-GTT CCC AAG TAC CCA AAT GAT GAG AAA CCA GT-3’ |
| Primer for mRNA sequencing-F | 5’-GTG ATG GGG CTG TGA GGC GTC-3’ |
| Primer for mRNA sequencing-R | 5’-TCC CCA TGT CAA AGT ATT CAG TGG GT-3’ |
| gRNA1 for CRISPR/Cas9 genome editing | 5’-CCA TCG TCA TTT CTT GAT TAA CG-3’ |
| gRNA2 for CRISPR/Cas9 genome editing | 5’-CAC GCT ACT GCA CTC CAA ACT GG-3’ |

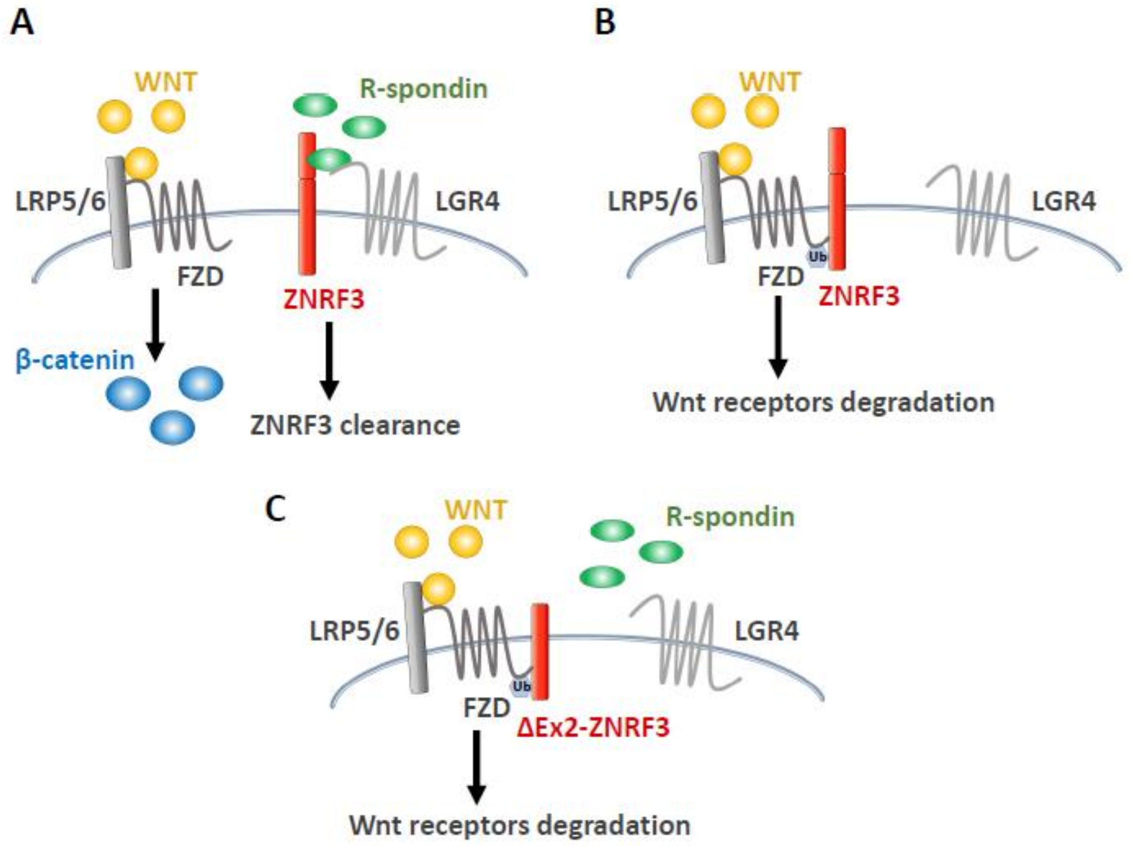


**Figure S1.**

ZNRF3 function on Wnt/β-catenin signaling. A. In the presence of R-spondins, R-spondins bind to both ZNRF3 and LGR4, leading to remove ZNRF3 and to stabilize Wnt receptors (Frizzled and LRP5/6) on the plasma membrane. Wnt can activate Wnt-β catenin signaling through binding to Wnt receptors. B. In the absence of R-spondins, ZNRF3 associates with Wnt receptors for ubiquitination of Frizzled, and promotes degradation of the Wnt receptors, resulting in attenuated Wnt-β catenin signaling. C. ΔEx2-ZNRF3 loses the ability of interaction with R-spondins and inactivates Wnt-β catenin signaling despite the presence of R-spondins. FZD and Ub denote Frizzled and ubiquitination, respectively.

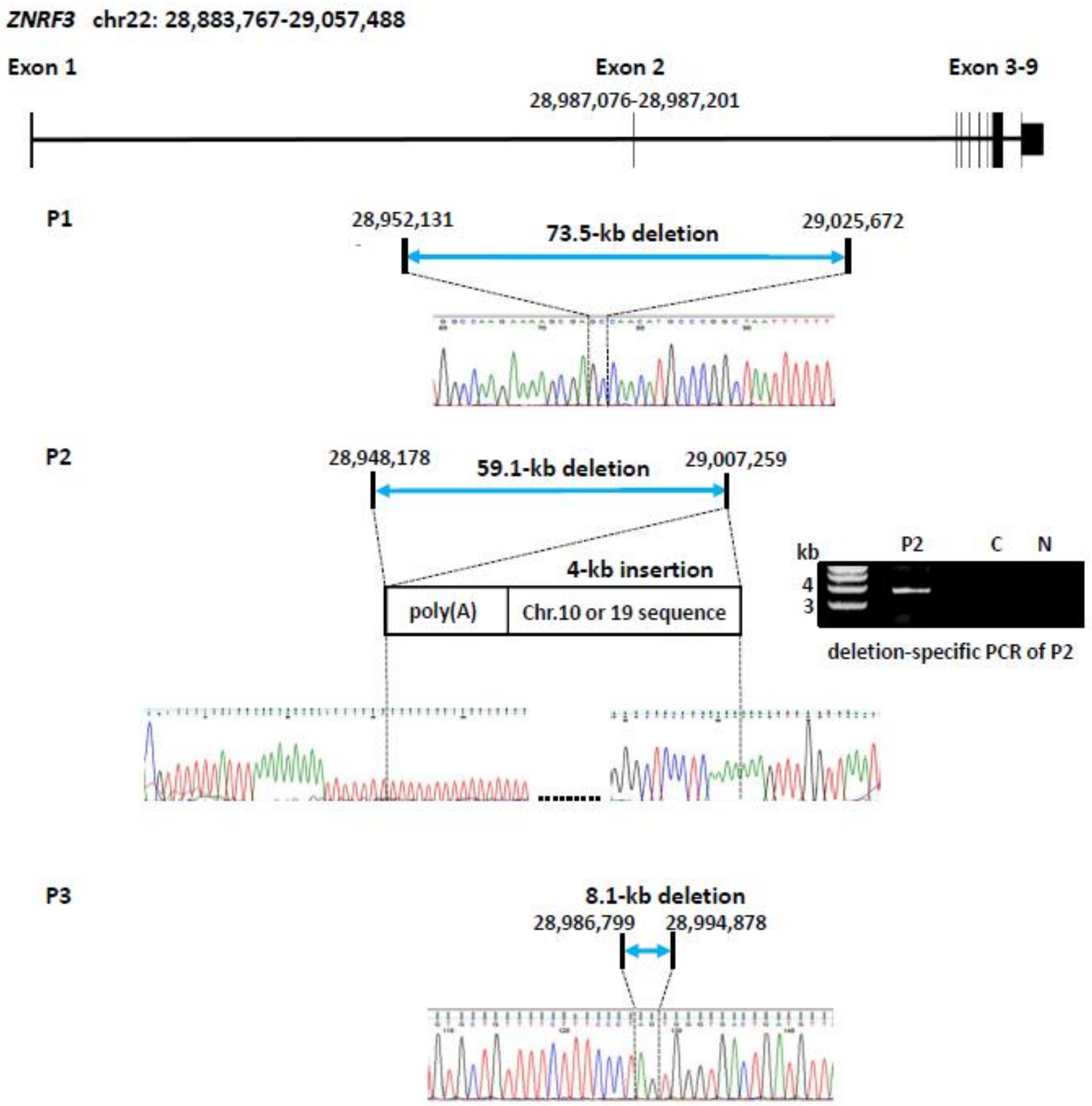


**Figure S2.**

Sequence chromatograms of the deletion breakpoints for P1-3. The gene structure of *ZNRF3* are shown on the top, and the localization of deletion breakpoints are mapped according to the human reference sequence GRCh37. P1 and P3 have simple deletions (73.5 kb and 8.1 kb) encompassing exon 2. P2 have 59.1-kb deletion and 4-kb insertion of poly (A) and Chr10 or 19 sequences between the breakpoints.

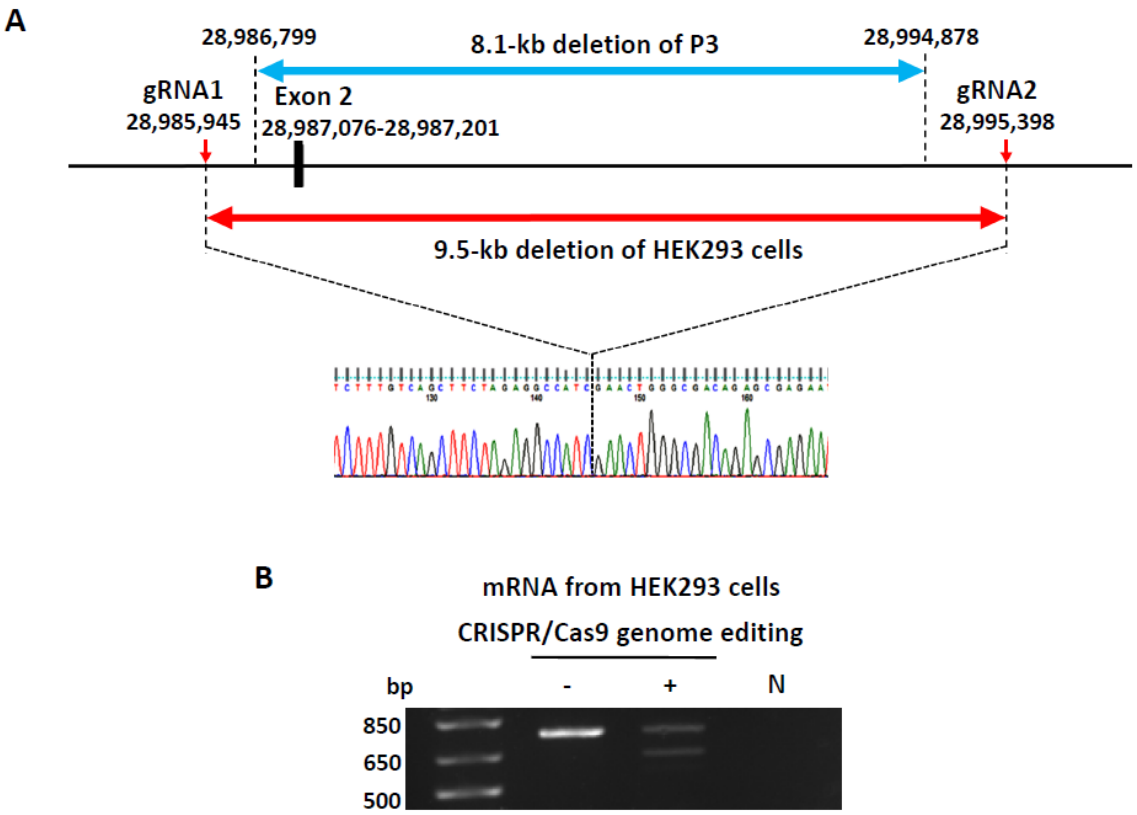


**Figure S3.**

A. Sequence chromatograms of the deletion breakpoints of the HEK293 cells with 9.5-kb deletion (hg19 Chr22: 29,381,933-29,391,386) generated using CRISPR/Cas9-based genome editing. The red arrows indicate the localization of gRNAs surrounding 8.1-kb deletion of P3. B. Agarose-gel electrophoresis of RT-PCR amplicons of mRNA from HEK293 cells using the primers spanning *ZNRF3* exons 1 to 5 (Figure 1A). HEK293 cells with CRISPR/Cas9-based genome editing have two fragments of 759-bp and 633-bp, as well as P1 and P2 (Figure 1D), and those without the genome editing has the only 759-bp fragment. We confirmed the presence of the identical 126-bp deleted sequence (data not shown).

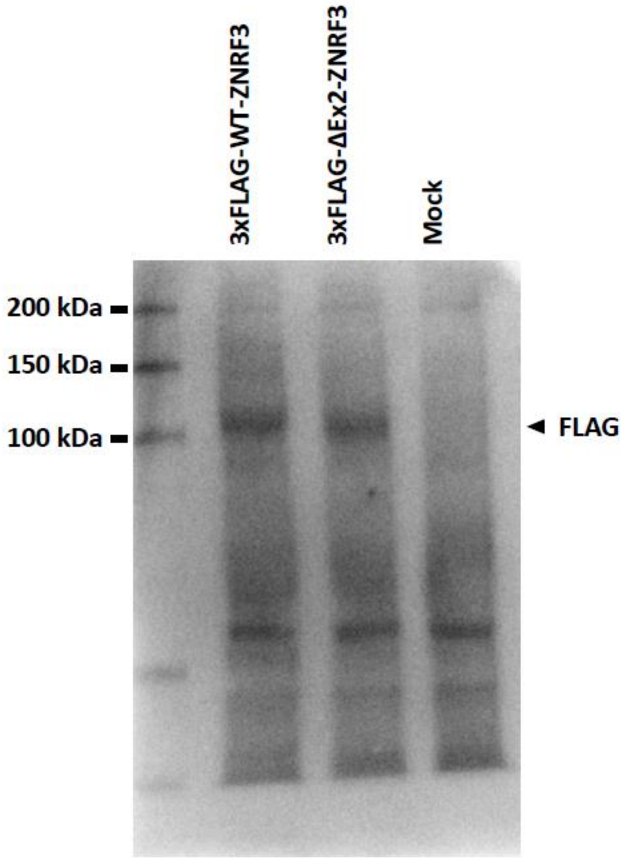


**Figure S4.**

Western blotting for the expression of ZNRF3 protein.
